# Estimating overdiagnosis in giant cell arteritis diagnostic pathways using genetic data: genetic association study

**DOI:** 10.1101/2023.04.17.23288682

**Authors:** Charikleia Chatzigeorgiou, Jennifer H Barrett, Javier Martin, UK GCA Consortium, Ann W Morgan, Sarah L Mackie

**Author notes:** Corresponding author, Leeds Institute of Cardiovascular and Metabolic Medicine, LIGHT Building, University of Leeds, Leeds, LS2 9JT.

## Abstract

**Objectives:** Prompt diagnosis of giant cell arteritis (GCA) is important to avert visual loss. False-negative temporal artery biopsy (TAB) can occur. Without vascular imaging, GCA may be overdiagnosed in TAB-negative cases, but it is unclear how often this occurs. An unbiased test is a way to address an imperfect reference standard. We used the known Human Leukocyte Antigen (HLA) region genetic association of TAB-positive GCA to estimate the extent of overdiagnosis before widespread adoption of temporal artery ultrasound as a first-line test.

**Methods:** Patients diagnosed with GCA between 1990-2014 consented to the UKGCA Consortium study. HLA region variants were jointly imputed from genome-wide genotypic data of cases and controls. Per-allele frequencies across all variants with p<1.0×10^−5^ were compared with population control data to estimate overdiagnosis rates in cases without a positive TAB.

**Results:** Genetic data from 663 patients diagnosed with GCA were compared with data from 2619 population controls. TAB-negative GCA (n=147) and GCA without a TAB result (n=160) had variant frequencies intermediate between those of TAB-positive GCA and population controls. Making several strong assumptions, we estimated that around two-thirds of TAB-negative cases and around one-third of cases without TAB result may have been overdiagnosed. From these data, TAB sensitivity is estimated at around 88%.

**Conclusions:** Conservatively assuming 95% specificity, TAB has a negative likelihood ratio of around 0.12. Genotyping alone cannot diagnose GCA at the individual level. Group-level HLA variant genotyping might be used to compare the overall accuracy of different diagnostic pathways or different classification criteria sets.

**Key messages:**

1. Under certain conditions and assumptions, overdiagnosis can be estimated using genetic data.
2. The specificity of temporal artery biopsy was estimated as about 88%.
3. Without vascular imaging, giant cell arteritis may often be overdiagnosed in biopsy-negative patients.

## Introduction

Giant cell arteritis (GCA) is a vasculitis of older people(1) which can present with a variety of symptoms including headache, scalp tenderness, jaw ache, ocular ischaemia, fever and weight loss(2). Where GCA is strongly suspected, it should be treated with high-dose glucocorticoids pending further investigation. The “diagnostic momentum” thus created can make it difficult to reverse the diagnosis if the temporal artery biopsy (TAB) subsequently proves negative and yet no better explanation for the presenting symptoms is found. Some patients with genuine GCA do have a negative TAB(3), often attributed to patchy arterial involvement or “skip lesions”(4, 5). Sensitivity of a unilateral TAB was estimated from bilateral TAB studies as 87%(6). Sometimes there is no evidence of any cranial artery involvement in GCA(7); this extracranial or “large-vessel” GCA subset is usually diagnosed with vascular imaging (8, 9). Vascular imaging is now also recommended as first-line investigation for cranial GCA, but where imaging and TAB give discordant or unexpected results, expert clinical judgement of GCA probability is crucial(10).

Where confirmatory tests are equivocal or negative, but pre-test probability is high, a clinical diagnosis of GCA may be justified by the desire to avoid preventable sight loss(11). The cost of this strategy is that some patients will be overdiagnosed. Adoption of better tests including vascular imaging should improve overall diagnostic accuracy of the pathway, but so far no method has been established that can demonstrate reduction in overdiagnosis.

We previously demonstrated that TAB-positive GCA is strongly associated with genetic variants in the Human Leucocyte Antigen (HLA) region including the single nucleotide polymorphisms rs9268905 (p=1.9×10^−54^, per-allele odds ratio(OR)=1.79) located between *HLA-DRA1* and *HLA-DRB1*, rs477515 (p=4.0×10^−40^, OR=1.73) located in the intergenic region between *HLA-DRB1* and *HLA-DQA1*, and the *HLA-DRB1*\*04 allele (p= 6.8×10^−38^, OR=1.92) (12). HLA region genotyping is not currently performed in routine care for GCA diagnosis; therefore genotype cannot directly influence diagnostic decisions and can act as an “unbiased umpire” (13). In this study, we sought to estimate the extent of GCA overdiagnosis during the era before widespread adoption of vascular imaging as a first-line test in GCA diagnostic pathways.

## Methods

### Study population

The UK GCA Consortium was designed as a genetic epidemiology study. Ethical approval was received from the Yorkshire and Humber–Leeds West Research Ethics Committee (REC Ref. 05/Q1108/28). Patients diagnosed with GCA by a consultant rheumatologist or ophthalmologist were identified from clinic lists and searches of TAB records from histopathology databases. Participants from 31 centres provided written informed consent. Clinical data and TAB result were determined by casenote review. TAB result was determined from the clinical records as “positive”, “negative” or “no result”. The “no result” category included cases where no biopsy was done, no temporal artery biopsy was done (no arterial tissue received for processing), or where no clear TAB result was determinable despite complete review of the patient’s medical records.

Population control data came from two sources: 1,430 individuals from the 1958 British Birth Cohort (58BC), who were born in England, Wales and Scotland during one week in 1958; and 1,500 healthy blood donors from the United Kingdom Blood Service for the Wellcome Trust Case Control Consortium (WTCCC)(14, 15) (Supplementary Methods).

The genetic data from the TAB-positive GCA cases have been previously included in two international collaborative studies(12, 16). In this study, we focus on the cases that received a “clinical diagnosis” of GCA: the TAB-negative and “no TAB result” cases that still received a GCA diagnosis.

### Genotyping and quality control

Genomic DNA was extracted from blood samples by standard methods(12). Cases and controls were genotyped separately with the use of two versions of the Illumina chip (Infinium Human core 24 Beadchip, HumanCore-12v1-0_A). The same quality filters were independently applied to the raw data from each cohort using PLINK version 1.9 (Supplementary Methods). The genome-wide Manhattan plot for GCA susceptibility is dominated by the signal from the HLA region in chromosome 6 (chr6: 29–34 Mb on build 36/hg18)(17) and therefore we chose to focus on the HLA region in our analysis. In this region, there is strong linkage disequilibrium. The allele type for each HLA gene can be inferred from SNP genotyping results by imputation.

### HLA imputation

Classical HLA alleles were determined from the single nucleotide polymorphism (SNP) data by imputation. Genotypic data from both cases and controls were jointly imputed across the extended major histocompatibility complex (MHC) using the SNP2HLA imputation method, using the Beagle software package (18, 19) (Supplementary Methods). Post-quality control filter thresholds were set to remove variants with a minor allele frequency (MAF) <0.01 and variants with an information score <0.8. After this was done, 159 classical HLA alleles and 6730 SNPs remained. Variants from the association analysis of TAB positive cases versus controls spanning the entire HLA region with p < 1.0 × 10^−5^ were used in order to estimate the misclassification rate.

### Association analysis using different case definitions

Firstly, to determine if the TAB result was associated with specific HLA alleles imputed across extended haplotypes, we compared the strength of association (odds ratio) and allele frequencies of susceptibility and protective HLA alleles in TAB-positive GCA, TAB-negative GCA and GCA with no TAB result.

### Estimation of misclassification rate

Because of the extensive linkage disequilibrium within the HLA region, different genetic variants within this region are not independent of one another, and so cannot be treated as independent variables. On the other hand, selection of only a few, very strong HLA associations of TAB-positive GCA would have potentially introduced “winner’s curse” bias (20). To address this, for each TAB-defined subset of the GCA cases, the proportion of patients misclassified as GCA was estimated by taking the average effect size across all the variants with p < 1.0 × 10^−5^ (per-allele association of positive-TAB GCA cases with controls). This conservative approach acknowledges the non-independence of alleles and SNPs from each other, while avoiding loss of potentially informative data. The proportions of “genuine” GCA cases in the TAB-negative GCA and GCA without TAB groups, p_n_ and p_x_, were calculated from the allele frequencies in TAB-positive cases and controls (Supplementary Methods). For simplicity, we assumed an additive gene-dose model in our analyses: although HLA-DRB1*04 may have a dominant effect (21) we could not assume this was also true of other HLA variants.

### Assumptions made

For the purposes of analysis we assumed (1) a 100% specificity of TAB (no false positive TAB), (2) the group of GCA cases without positive TAB comprises a mixture of “genuine” GCA and “overdiagnosed” GCA, (3) all “genuine” GCA cases share the same HLA associations, (4) “overdiagnosed” cases have a similar distribution of HLA variants as the control population, (5) HLA status did not influence clinical diagnosis.

## Results

After quality control, we had genetic data for 663 cases of GCA and 2619 population controls. The GCA patients were diagnosed between 1991 and 2014. Only 65 GCA patients had imaging evidence of GCA: during this period, TAB was the first-line confirmatory test, and vascular imaging tests for GCA were not routinely performed in most UK hospitals. Table 1 gives clinical data on the TAB-positive GCA (n=356), TAB-negative GCA (n=147) and cases of GCA without any TAB result (n=160).

**Table 1.**
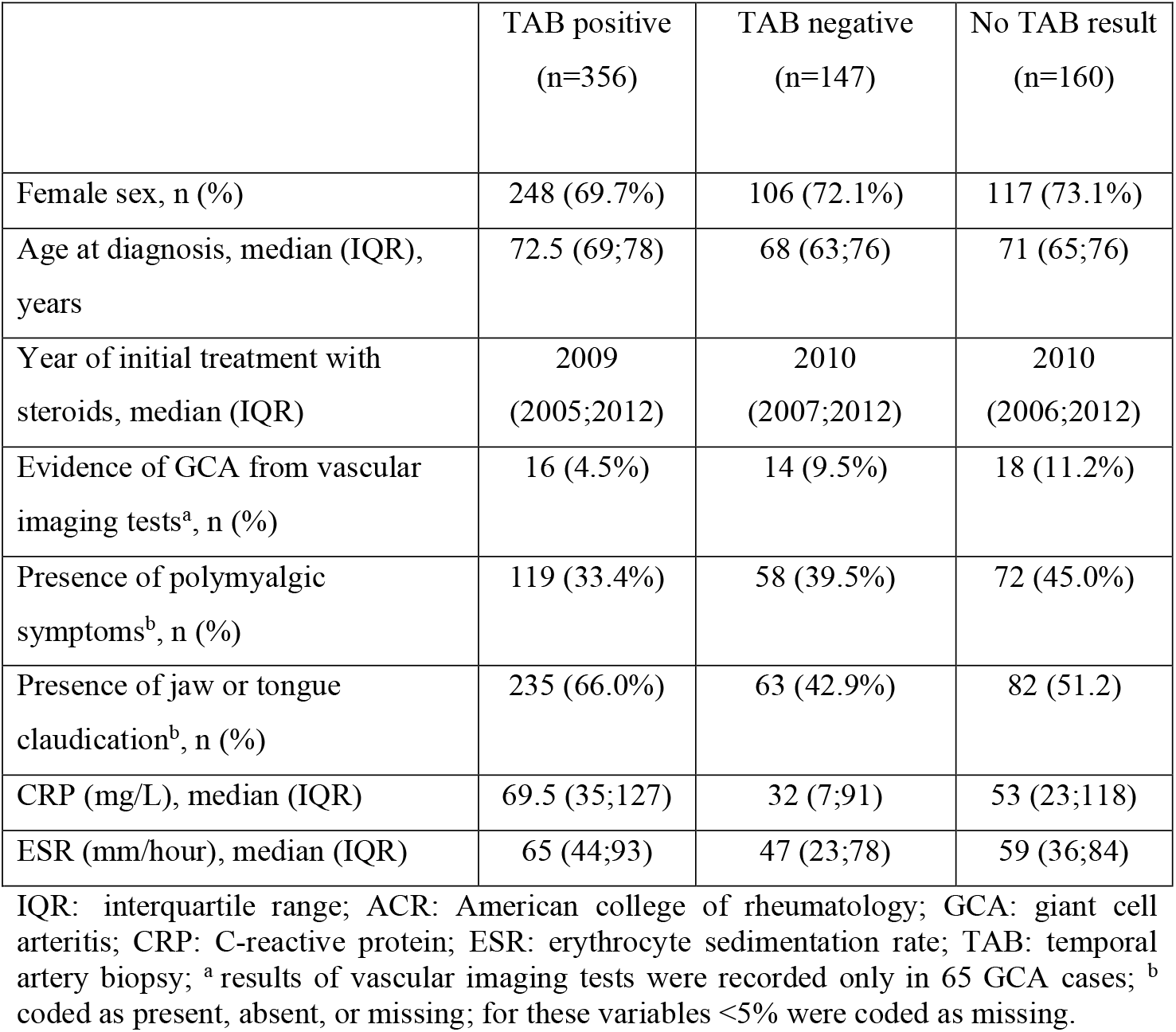
Clinical features of GCA patients included in the analysis

|  | TAB positive<br>(n=356) | TAB negative<br>(n=147) | No TAB result<br>(n=160) |
| --- | --- | --- | --- |
| Female sex, n (%) | 248 (69.7%) | 106 (72.1%) | 117 (73.1%) |
| Age at diagnosis, median (IQR), years | 72.5 (69;78) | 68 (63;76) | 71 (65;76) |
| Year of initial treatment with steroids, median (IQR) | 2009<br>(2005;2012) | 2010<br>(2007;2012) | 2010<br>(2006;2012) |
| Evidence of GCA from vascular imaging tests<sup>a</sup>, n (%) | 16 (4.5%) | 14 (9.5%) | 18 (11.2%) |
| Presence of polymyalgic symptoms<sup>b</sup>, n (%) | 119 (33.4%) | 58 (39.5%) | 72 (45.0%) |
| Presence of jaw or tongue claudication<sup>b</sup>, n (%) | 235 (66.0%) | 63 (42.9%) | 82 (51.2) |
| CRP (mg/L), median (IQR) | 69.5 (35;127) | 32 (7;91) | 53 (23;118) |
| ESR (mm/hour), median (IQR) | 65 (44;93) | 47 (23;78) | 59 (36;84) |

IQR: interquartile range; ACR: American college of rheumatology; GCA: giant cell arteritis; CRP: C-reactive protein; ESR: erythrocyte sedimentation rate; TAB: temporal artery biopsy; <sup>a</sup> results of vascular imaging tests were recorded only in 65 GCA cases; <sup>b</sup> coded as present, absent, or missing; for these variables <5% were coded as missing.

### HLA allele associations

The HLA association of TAB-positive GCA patients was similar to that described in previous studies, including the large international study to which we had contributed TAB-positive cases(12). *HLA-DQA1*\*01, *HLA-DQB1*\*05 and *HLA-DQB1*\*06 alleles had a protective effect; *HLA-DQA1*\*03, *HLA-DQB1*\*03 and *HLA-DRB1*\*04 were associated with GCA susceptibility (Table 2, “TAB positive” column).

**Table 2.** Significant association of classical HLA alleles selected from the TAB positive cohort, at two- and four-digit resolution, and then tested in the subsets of negative and no TAB result at 0.00001 level of significance without conditioning

|  |  | Positive TAB (n=356)<br>vs controls (n=2619) |  |  | Negative TAB (n=147)<br>vs controls (n=2619) |  |  | No TAB result (n=160)<br>vs controls (n=2619) |  |  |
| --- | --- | --- | --- | --- | --- | --- | --- | --- | --- | --- |
| Classical HLA allele | MAF<sub>ctrl</sub> | MAF<sub>pos</sub> | OR<sub>pos</sub> | P<sub>pos</sub> | MAF<sub>neg</sub> | OR<sub>neg</sub> | P<sub>neg</sub> | MAF<sub>unknown</sub> | OR<sub>unknown</sub> | P<sub>unknown</sub> |
| HLA-DQA1*01 | 0.38 | 0.23 | 0.50 | 8.07 x 10<sup>-14</sup> | 0.34 | 0.83 | 0.14 | 0.28 | 0.62 | 0.0001 |
| HLA-DQA1*0102 | 0.19 | 0.12 | 0.57 | 5.43 x 10<sup>-06</sup> | 0.16 | 0.80 | 0.18 | 0.11 | 0.53 | 0.0006 |
| HLA-DQA1*03 | 0.21 | 0.33 | 1.85 | 2.37 x 10<sup>-12</sup> | 0.28 | 1.42 | 0.0086 | 0.29 | 1.49 | 0.0020 |
| HLA-DQA1*0301 | 0.21 | 0.33 | 1.85 | 2.37 x 10<sup>-12</sup> | 0.28 | 1.42 | 0.0086 | 0.29 | 1.49 | 0.0020 |
| HLA-DQB1*03 | 0.35 | 0.49 | 1.76 | 2.65 x 10<sup>-12</sup> | 0.41 | 1.28 | 0.04 | 0.41 | 1.29 | 0.0282 |
| HLA-DQB1*0301 | 0.18 | 0.25 | 1.46 | 6.00 x 10<sup>-4</sup> | 0.21 | 1.17 | 0.28 | 0.21 | 1.18 | 0.24 |
| HLA-DQB1*0302 | 0.11 | 0.18 | 1.83 | 1.59 x 10<sup>-08</sup> | 0.17 | 1.62 | 0.0029 | 0.17 | 1.65 | 0.0013 |
| HLA-DQB1*05 | 0.15 | 0.08 | 0.52 | 4.75 x10<sup>-06</sup> | 0.14 | 0.95 | 0.79 | 0.13 | 0.84 | 0.32 |
| HLA-DQB1*06 | 0.23 | 0.15 | 0.58 | 7.68 x 10<sup>-07</sup> | 0.20 | 0.80 | 0.15 | 0.15 | 0.56 | 0.0004 |
| HLA-DRB1*04 | 0.20 | 0.32 | 1.96 | 5.13 x 10<sup>-14</sup> | 0.27 | 1.48 | 0.0038 | 0.29 | 1.65 | 0.0001 |
| HLA-DRB1*0401 | 0.12 | 0.20 | 1.94 | 1.54 x 10<sup>-09</sup> | 0.14 | 1.28 | 0.17 | 0.17 | 1.65 | 0.0017 |
| HLA-DRB1*0404 | 0.05 | 0.09 | 2.16 | 4.97 x 10<sup>-07</sup> | 0.08 | 1.77 | 0.019 | 0.07 | 1.64 | 0.038 |

P<sub>-</sub>: p<sub>value</sub> from the logistic regression of each subgroup of cases (positive, negative, unknown) with the control individuals, OR: per-allele odds ratio assuming an additive model, MAF: minor allele frequency

TAB-negative GCA showed a pattern of HLA allelic associations in the same direction as TAB-positive GCA, but the effect was generally diluted: for each HLA allele, the strength of association was diminished (closer to the null odds ratio of 1.0). The odds ratios for GCA with no biopsy result were generally intermediate between those for TAB-positive and TAB-negative GCA (Table 2: columns for TAB negative, no TAB result).

### Single-nucleotide polymorphisms

Associations with the three SNPs (rs9268969, rs9275184, rs477515) previously reported as being associated with biopsy-confirmed GCA (12, 16) were markedly diluted for TAB-negative GCA, to the extent that only rs9275184 (located between? *HLA-DQA1* and *HLA-DQA2)* retained statistical significance at p<0.05 (Table 3, “negative TAB” column). Again, odds ratios for GCA with no biopsy result were intermediate between those observed for TAB-positive GCA and TAB-negative GCA (Table 3, “no TAB result” column).

**Table 3.** Previously-reported independent SNP associations in the HLA region among subsets of GCA classified according to TAB result [12, 16].

|  |  |  |  | All cases (n=663)<br>vs controls (n=2619) |  |  | Positive TAB (n=356)<br>vs controls (n=2619) |  |  | Negative TAB (n=147)<br>vs controls (n=2619) |  |  | No TAB result (n=160)<br>vs controls (n=2619) |  |  |
| --- | --- | --- | --- | --- | --- | --- | --- | --- | --- | --- | --- | --- | --- | --- | --- |
| SNP | BP | Gene/<br>Intergenic<br>region | MA<br>F<sub>CTR</sub><br>L | MAF<sub>a</sub><br>ll | OR<sub>al</sub><br>l | P<sub>all</sub> | MAF<sub>pos</sub> | OR<sub>pos</sub> | P<sub>pos</sub> | MAF<sub>neg</sub> | OR<sub>neg</sub> | P<sub>neg</sub> | MAF<sub>unk</sub> | OR<sub>unk</sub> | P<sub>unk</sub> |
| rs477515 | 32677668 | *HLA-DRB1*<br>/*HLA-DQA1* | 0.35 | 0.46 | 1.54 | $2.99 \times 10^{-12}$ | 0.50 | 1.84 | $4.14 \times 10^{-14}$ | 0.37 | 1.09 | 0.49 | 0.44 | 1.43 | 0.002 |
| rs9268969 | 32542328 | *HLA-DRA1*<br>/*HLA-DRB1* | 0.37 | 0.47 | 1.49 | $1.99 \times 10^{-10}$ | 0.51 | 1.78 | $1.03 \times 10^{-12}$ | 0.38 | 1.06 | 0.61 | 0.44 | 1.34 | 0.01 |
| rs9275184 | 32762692 | *HLA-DQA1*<br>/*HLA-DQA2* | 0.11 | 0.18 | 1.75 | $5.48 \times 10^{-11}$ | 0.18 | 1.83 | $1.53 \times 10^{-08}$ | 0.17 | 1.62 | 0.003 | 0.17 | 1.64 | 0.001 |

BP: Base pair position; MAF: minor allele frequency; OR: odds ratio

### Estimation of misclassification rates

470 HLA variants were associated with TAB-positive GCA compared to controls at a per-allele threshold of p<1.0 × 10^−5^. We estimated the proportion p_n_ of “genuine” cases among TAB-negative GCA as 0.33 (SD:0.23) and proportion p_x_ of “genuine” cases in GCA without TAB result as 0.67 (SD:0.15).

## Discussion

In this study, we used HLA genotyping data to estimate the proportion of clinically-diagnosed GCA patients who are overdiagnosed. Of 503 cases in our study who had a TAB, 356 (71%) were recorded as positive; this is in line with the 77% sensitivity of TAB reported in a meta-analysis(22). We found, by demonstrating dilution of the known genetic associations with biopsy-positive GCA, that subject to strong assumptions, about two-thirds of biopsy-negative GCA cases, and one-third of GCA cases without a biopsy result, may have been overdiagnosed.

The Merriam-Webster dictionary defines overdiagnosis as “the diagnosis of a condition or disease more often than it is actually present”. The problem here is that even if it is shown at the group level that overdiagnosis is occurring, overdiagnosis/overtreatment of an individual patient remains a rational response to diagnostic uncertainty, if the potential consequences of missing a case of genuine GCA include significant adverse outcomes such as visual loss. Under such circumstances, the greater the diagnostic uncertainty, the more overdiagnosis is likely to occur.

Our estimate depends on several assumptions which are limitations of our approach. **(i)** We assumed a 100% specificity of TAB (no false positive TAB). This assumption has been called into question by a reliability study in which discordant classifications of TAB images as positive/negative were made by a panel of pathologists(23). Standardised reporting templates for TAB might address this. **(ii)** We assumed that the group of GCA cases without positive TAB comprises a mixture of “genuine” GCA and “overdiagnosed” GCA. This could be an oversimplification if there are “halfway-house” disease states including potentially PMR.(9) **(iii)** We assumed that all “genuine” GCA cases share the same HLA associations, but there may be heterogeneity within this disease. Large-vessel involvement was previously suggested in small studies to have a different pattern of HLA association (7), but a more recent study had failed to confirm this(24). Takayasu arteritis (TAK) does appear to have a different HLA association compared to GCA (17, 25), but the cases in our study were all of an age-range compatible with GCA rather than TAK. **(iv)** We assumed “overdiagnosed” cases have a similar distribution of HLA variants to population controls. Although there is global variation of population HLA frequencies (21), our cases and controls both came from the UK. **(v)** We assumed that HLA genotype did not influence clinical diagnosis. This assumptions seems reasonable, since the clinical diagnosis of GCA was an inclusion criterion for UK GCA Consortium, and HLA typing is not an accepted test for GCA diagnosis in clinical practice. The HLA genotyping done for this study was only done in retrospect and the results of the HLA genotyping were not returned to the clinical care team.

Overdiagnosis can be a system-level phenomenon rather than necessarily implying error on the diagnostician’s part(26). Given an estimated GCA prevalence of 52 per 100,000 in the over-50s(1), with just over 25 million over-50s living in the UK (2021 data (27)), over 13,000 people in the UK have ever been diagnosed with GCA. Extrapolating from our findings reported here, up to 3,500 of these people may have been overdiagnosed and therefore been exposed to the harms of long-term glucocorticoid therapy. However, diagnostic pathways have rapidly evolved since the 2007-2014 era.

Although 71% of GCA patients in our cohort had a positive TAB, removing “overdiagnosed” cases from the denominator would give a TAB sensitivity for detecting “genuine GCA” of around 88%, comparable to the 87% sensitivity reported by studies of bilateral TAB(6). For the diagnostician, negative likelihood ratio (ratio of pre-test odds to post-test odds, given a negative test result) is of more practical use. Taking this 88% sensitivity and conservatively assuming a TAB specificity of 95%, the negative likelihood ratio associated with a negative TAB would be 0.12 (revised downward from the previously-reported estimate of 0.23(28)). To give an illustrative example, pre-test GCA probability of 20%, 50% or 80% would be downgraded by a negative TAB result to a post-test probability of 3%, 11% or 34% respectively.

Could HLA genotyping be useful in clinical practice to improve accuracy of estimation of pre-test probability? *HLA-DRB1*\*04, the strongest allelic association at the group level, is fairly common in northern European populations and having this allele would produce a comparatively small diagnostic shift at the individual patient level: the positive and negative likelihood ratios of *HLA-DRB1*\*04 in a northern European population are approximately 1.79 and 0.76 respectively (24); this could be combined with likelihood ratios for symptoms, signs and laboratory markers of inflammation to incrementally improve diagnostic accuracy(2). It is conceivable that HLA genetic data might be incorporated into a future multiparameter clinical decision aid, most likely, including just one or a few HLA variants into a model would be more feasible than analysing all 470 HLA variants we studied here. Because of strong linkage disequilibrium and multiplicity of testing, it is not possible to identify the optimal variant(s) from our data.

Our results provide a benchmark for estimating GCA overdiagnosis rates during the era before widespread adoption of temporal artery ultrasound. Future studies of HLA frequencies in GCA cohorts diagnosed via pathways involving vascular imaging could ascertain whether diagnostic accuracy has improved with contemporary diagnostic pathways. Within cohorts diagnosed during the same time period, the accuracy of different classification criteria for GCA could also be compared; HLA variant/allele frequencies in the subgroup classified as GCA by the 1990 ACR criteria, might be compared to HLA variant/allele frequencies in the subgroup classified as GCA by the 2022 ACR/EULAR classification criteria. Unbiased tests have a useful role to play in such comparisons(13).

Lastly, our approach might be extended to evaluate diagnostic pathways or classification criteria for other diseases where the gold-standard test is highly specific but insensitive, and where a strong genetic association is present.

## Supporting information

Appendix 1 UK GCA Consortium Members

## Data Availability

The data used to support the findings of this study are included within the article.

## Acknowledgements

We would like to thank UK GCA participants, without whom this study would not have been possible.

## Author Contributions

Design, concept: CC, AWM, SLM, JHB

Analysis: CC, JHB, AWM, SLM

Clinical expertise: AWM, SLM

Writing manuscript: CC, AWM, SLM, JHB, JM

Manuscript review: CC, AWM, SLM, JHB, JM

## Data availabilite statement

The data used to support the findings of this study are included within the article.

## References

1. Li KJ, Semenov D, Turk M, Pope J. A meta-analysis of the epidemiology of giant cell arteritis across time and space. Arthritis research & therapy. 2021;23(1):82.

2. van der Geest KSM, Sandovici M, Brouwer E, Mackie SL. Diagnostic Accuracy of Symptoms, Physical Signs, and Laboratory Tests for Giant Cell Arteritis: A Systematic Review and Meta-analysis. JAMA Intern Med. 2020;180(10):1295–304.

3. Garvey TD, Koster MJ, Warrington KJ. My Treatment Approach to Giant Cell Arteritis. Mayo Clin Proc. 2021;96(6):1530–45.

4. Poller DN, van Wyk Q, Jeffrey MJ. The importance of skip lesions in temporal arteritis. J Clin Pathol. 2000;53(2):137–9.

5. Hernandez-Rodriguez J, Murgia G, Villar I, Campo E, Mackie SL, Chakrabarty A, et al. Description and Validation of Histological Patterns and Proposal of a Dynamic Model of Inflammatory Infiltration in Giant-cell Arteritis. Medicine (Baltimore). 2016;95(8):e2368.

6. Niederkohr RD, Levin LA. A Bayesian analysis of the true sensitivity of a temporal artery biopsy. Invest Ophthalmol Vis Sci. 2007;48(2):675–80.

7. Brack A, Martinez-Taboada V, Stanson A, Goronzy JJ, Weyand CM. Disease pattern in cranial and large-vessel giant cell arteritis. Arthritis Rheum. 1999;42:311–7.

8. van der Geest KSM, Sandovici M, van Sleen Y, Sanders JS, Bos NA, Abdulahad WH, et al. Review: What Is the Current Evidence for Disease Subsets in Giant Cell Arteritis? Arthritis & rheumatology. 2018;70(9):1366–76.

9. Dejaco C, Duftner C, Buttgereit F, Matteson EL, Dasgupta B. The spectrum of giant cell arteritis and polymyalgia rheumatica: revisiting the concept of the disease. Rheumatology. 2017;56(4):506–15.

10. Hellmich B, Agueda A, Monti S, Buttgereit F, de Boysson H, Brouwer E, et al. 2018 Update of the EULAR recommendations for the management of large vessel vasculitis. Annals of the rheumatic diseases. 2020;79(1):19–30.

11. Dasgupta B, Borg FA, Hassan N, Alexander L, Barraclough K, Bourke B, et al. BSR and BHPR guidelines for the management of giant cell arteritis. Rheumatology. 2010;49(8):1594–7.

12. Carmona FD, Mackie SL, Martin JE, Taylor JC, Vaglio A, Eyre S, et al. A large-scale genetic analysis reveals a strong contribution of the HLA class II region to giant cell arteritis susceptibility. American journal of human genetics. 2015;96(4):565–80.

13. Glasziou P, Irwig L, Deeks JJ. When should a new test become the current reference standard? Annals of internal medicine. 2008;149(11):816–22.

14. Wellcome Trust Case Control C, Australo-Anglo-American Spondylitis C, Burton PR, Clayton DG, Cardon LR, Craddock N, et al. Association scan of 14,500 nonsynonymous SNPs in four diseases identifies autoimmunity variants. Nat Genet. 2007;39(11):1329–37.

15. Wellcome Trust Case Control C. Genome-wide association study of 14,000 cases of seven common diseases and 3,000 shared controls. Nature. 2007;447(7145):661–78.

16. Carmona FD, Vaglio A, Mackie SL, Hernandez-Rodriguez J, Monach PA, Castaneda S, et al. A Genome-wide Association Study Identifies Risk Alleles in Plasminogen and P4HA2 Associated with Giant Cell Arteritis. American journal of human genetics. 2017;100(1):64–74.

17. Carmona FD, Coit P, Saruhan-Direskeneli G, Hernandez-Rodriguez J, Cid MC, Solans R, et al. Analysis of the common genetic component of large-vessel vasculitides through a meta-Immunochip strategy. Sci Rep. 2017;7:43953.

18. Jia X, Han B, Onengut-Gumuscu S, Chen WM, Concannon PJ, Rich SS, et al. Imputing amino acid polymorphisms in human leukocyte antigens. PLoS One. 2013;8(6):e64683.

19. Browning BL, Browning SR. A unified approach to genotype imputation and haplotype-phase inference for large data sets of trios and unrelated individuals. American journal of human genetics. 2009;84(2):210–23.

20. Wang WY, Barratt BJ, Clayton DG, Todd JA. Genome-wide association studies: theoretical and practical concerns. Nat Rev Genet. 2005;6(2):109–18.

21. Mackie SL, Taylor JC, Haroon-Rashid L, Martin S, Dasgupta B, Gough A, et al. Association of HLA-DRB1 amino acid residues with giant cell arteritis: genetic association study, meta-analysis and geo-epidemiological investigation. Arthritis research & therapy. 2015;17:195.

22. Rubenstein E, Maldini C, Gonzalez-Chiappe S, Chevret S, Mahr A. Sensitivity of temporal artery biopsy in the diagnosis of giant cell arteritis: a systematic literature review and meta-analysis. Rheumatology. 2020;59(5):1011–20.

23. Luqmani R, Lee E, Singh S, Gillett M, Schmidt WA, Bradburn M, et al. The Role of Ultrasound Compared to Biopsy of Temporal Arteries in the Diagnosis and Treatment of Giant Cell Arteritis (TABUL): a diagnostic accuracy and cost-effectiveness study. Health Technol Assess. 2016;20(90):1–238.

24. Prieto-Pena D, Remuzgo-Martinez S, Ocejo-Vinyals JG, Atienza-Mateo B, Munoz-Jimenez A, Ortiz-Sanjuan F, et al. Cranial and extracranial giant cell arteritis share similar HLA-DRB1 association. Seminars in arthritis and rheumatism. 2020;50(5):897–901.

25. Ortiz-Fernandez L, Saruhan-Direskeneli G, Alibaz-Oner F, Kaymaz-Tahra S, Coit P, Kong X, et al. Identification of susceptibility loci for Takayasu arteritis through a large multi-ancestral genome-wide association study. American journal of human genetics. 2021;108(1):84–99.

26. Brodersen J, Kramer BS, Macdonald H, Schwartz LM, Woloshin S. Focusing on overdiagnosis as a driver of too much medicine. BMJ. 2018;362:k3494.

27. Overview of the UK population: January 2021 2021 [

28. Mackie SL, Brouwer E. What can negative temporal artery biopsies tell us? Rheumatology. 2020;59(5):925–7.

