## Supplementary Methods for "Estimating overdiagnosis in giant cell arteritis diagnostic pathways using genetic data: genetic association study"

### Case-control cohorts

*UK GCA consortium*

All cases were of European descent; they were recruited from 32 hospital sites across the UK between 2005 and 2009 [1, 2]. Individuals from this study who had provided informed written consent to participate in genetic studies and on whom genetic data were available on 17/02/2017 were included in this analysis.

*Wellcome Trust Case Control Consortium controls*

The control individuals came from two sources: 1,430 individuals from the 1958 British Birth Cohort (58BC) including individuals born in England, Wales and Scotland during one week in 1958, and 1,500 healthy blood donors recruited from the United Kingdom Blood Service (UK Blood Services, UKBS, controls) for the Wellcome Trust Case Control Consortium (WTCCC) [3, 4].

### Genotyping and Quality control

Genomic DNA from peripheral blood samples of all GCA cases was extracted using the Genecatcher method (company name: Thermo Fisher Scientific), and sent to the Genomics and Genotyping Unit of the Pfizer-University of Granada-Junta de Andalucía Centre for Genomics and Oncological Research (GENYO, Granada, Spain, *Batch 1* and *Batch 2*) according to the manufacturer's protocol in order to be analysed with the use of the GWAS platform ''Infinium HumanCore Beadchip'' in an iScan System and the Genotyping Module (v.1.9) of the GenomeStudio software (Illumina). Cases were genotyped in two slightly different batches of the same chip (Infinium Human core 24 Beadchip, HumanCore-12v1-0_A and HumanCore-24v1). For the control group (*Batch 3*), genotyping was performed using the Illumina 1.2M (custom) chip (Illumina Human, 12 Beadchip) [3].

Data from all the different batches were filtered separately based on the same analytical protocol using PLINK v.1.9 [5]. SNPs with a call rate <0.97, a significant departure from Hardy-

Weinberg equilibrium (HWE) ($p<10^{-8}$ in cases or $p<10^{-5}$ in controls), or a minor allele frequency (MAF) <0.01 were removed. Based on the SNPs that passed the above quality control (QC) thresholds, samples were then excluded if they had a call rate <97%, or if there were inconsistencies between recorded and genotype-inferred gender or excess heterozygosity on the autosomes. Duplicates and first-degree or second-degree relatives were identified based on identity-by-descent statistics and the individuals with the lower genotype rate were removed. Genetic data from cases and controls were merged separately with HapMap data and a principal component analysis (PCA) was performed with the use of Plink software version 1.9. Individuals were excluded from this study if they were found to deviate from the European Hapmap cluster.

HLA imputation

The markers spanning 29–34 Mb (GRCh36) on chromosome 6, which encompassed the HLA region, were extracted from the post-QC dataset. All cases (regardless of TAB outcome) genotyped in Spain (*Batch* 1 and *Batch* 2) and controls were imputed together using SNP2HLA (V.1.0) (http://www.broadinstitute.org/ mpg/snp2hla/) [6]. This is an approach which enables imputation of classical HLA two and four-digit alleles as well as individual amino acids encoded by these HLA alleles, which may play an important role in protein function. Knowledge of the patterns of linkage disequilibrium (LD) enabled the inference of HLA alleles, amino acids and SNPs across the whole HLA region. The method uses a large reference dataset collected by the Type 1 Diabetes Genetics consortium (T1DGC) (n=5225) [7]. This dataset has gold-standard HLA typing providing high-density SNP data; the imputation uses the Beagle software package [6] and has been previously used by a number of researchers [8-10].

Association analysis using different case definitions

The results of the association analysis are presented for previously associated variants for TAB-positive confirmed GCA cases [11, 12] (rs9268905, rs9275592, rs477515 and HLA-DRB1*04). In practice, rs9268969 was analysed as a proxy for rs9268905 and rs9275184 as a proxy for rs9275592, as rs9268905 and rs9275592 were not included in the imputed data (rs9268969 and rs9268905, rs9275592 and rs9275184 are in complete LD, RSquare:1.0,

DPrime:1.0). This is because a different reference panel was used in the recent GWAS paper (1000 Genome Project Phase III) to the reference panel that was used in this study (Type 1 Diabetes Mellitus Genetics Consortium) [11]. According to the results from the SNP Annotation and Proxy (SNAP) search from the Broad Institute (http://archive.broadinstitute.org/mpg/snap/ldsearchpw.php), rs477515 and rs9268969 are in strong LD (RSquare:0.9, DPrime:0.8) and similar LD was observed for rs477515 and rs9268905. Variants rs9268905 and rs9268969 were found to be in complete LD (RSquare:1.0, DPrime:1.0).

Estimation of misclassification rates using allele frequencies

For the purpose of this analysis, 663 cases from the UK GCA Consortium (356 with a positive TAB, 160 with unknown TAB outcome and 147 with negative TAB outcome) and 2619 controls from the WTCCC were used. A total of 8044 variants in the HLA region were available after QC. The results from the logistic regression of i) positive TAB cases in comparison with controls, ii) negative TAB cases in comparison with controls and iii) unknown TAB in comparison with controls were merged in a single dataset including only the common variants (MAF >0.1 in controls) that had a p-value $<1.0 \times 10^{-5}$ from the association analysis of TAB positive cases versus controls.

For each of these variants, the observed allele frequencies in positive TAB cases ($f_1$), controls ($f_0$), negative TAB cases ($f_n$) and cases with an unknown TAB ($f_x$) were obtained. Let $p_n$ be the proportion of "true" cases among the group of cases with a negative TAB. Under the assumption that this group is actually a mixture of cases and controls, the expected value of the allele frequency $f_n$ in this group is given by:

$$p_n f_1 + (1 - p_n) f_0$$

Similarly, if $p_x$ is the proportion of true cases among the group of cases with unknown TAB, then the expected value of the allele frequency $f_x$ in this group is:

$$p_x f_1 + (1 - p_x) f_0$$

Given the observed allele frequencies in the different groups, these relationships were used to estimate $p_n$ and $p_x$ for each variant. To obtain an overall estimate of the proportion of cases correctly classified in the TAB negative subgroup, the mean of the estimates of $p_n$ across all variants was used. Similarly, for the subgroup with unknown TAB, the proportion was estimated by the mean of the variant-specific values of $p_x$.

References


1. Mackie, S.L., et al., *Association of HLA-DRB1 amino acid residues with giant cell arteritis: genetic association study, meta-analysis and geo-epidemiological investigation.* Arthritis Res Ther, 2015. **17**: p. 195.
2. Mackie, S.L., et al., *Ischaemic manifestations in giant cell arteritis are associated with area level socio-economic deprivation, but not cardiovascular risk factors.* Rheumatology (Oxford), 2011. **50**(11): p. 2014-22.
3. Wellcome Trust Case Control, C., et al., *Association scan of 14,500 nonsynonymous SNPs in four diseases identifies autoimmunity variants.* Nat Genet, 2007. **39**(11): p. 1329-37.
4. Wellcome Trust Case Control, C., *Genome-wide association study of 14,000 cases of seven common diseases and 3,000 shared controls.* Nature, 2007. **447**(7145): p. 661-78.
5. Purcell, S., et al., *PLINK: a tool set for whole-genome association and population-based linkage analyses.* Am J Hum Genet, 2007. **81**(3): p. 559-75.
6. Jia, X., et al., *Imputing amino acid polymorphisms in human leukocyte antigens.* PLoS One, 2013. **8**(6): p. e64683.
7. Rich, S.S., et al., *Overview of the Type I Diabetes Genetics Consortium.* Genes Immun, 2009. **10 Suppl 1**: p. S1-4.
8. Raychaudhuri, S., et al., *Five amino acids in three HLA proteins explain most of the association between MHC and seropositive rheumatoid arthritis.* Nat Genet, 2012. **44**(3): p. 291-6.
9. Han, B., et al., *Fine mapping seronegative and seropositive rheumatoid arthritis to shared and distinct HLA alleles by adjusting for the effects of heterogeneity.* Am J Hum Genet, 2014. **94**(4): p. 522-32.
10. Hu, X., et al., *Additive and interaction effects at three amino acid positions in HLA-DQ and HLA-DR molecules drive type 1 diabetes risk.* Nat Genet, 2015. **47**(8): p. 898-905.
11. Carmona, F.D., et al., *A Genome-wide Association Study Identifies Risk Alleles in Plasminogen and P4HA2 Associated with Giant Cell Arteritis.* Am J Hum Genet, 2017. **100**(1): p. 64-74.
12. Carmona, F.D., et al., *A large-scale genetic analysis reveals a strong contribution of the HLA class II region to giant cell arteritis susceptibility.* Am J Hum Genet, 2015. **96**(4): p. 565-80.
