## Appendix 1 UK GCA Consortium Members for "Estimating overdiagnosis in giant cell arteritis diagnostic pathways using genetic data: genetic association study"


**UK GCA Consortium**

**Management Team:** Ann W Morgan[1], Sarah L Mackie[1], Louise Sorensen[1]

**Laboratory Team:** Lubna Haroon Raashid[1], Steve Martin[1], James I Robinson[1], Sam Mellen[6], Sarah Hoggart[6],

**Analysis Team:** Jennifer H Barrett[1], John C Taylor[1]

**Consultants:** Colin Pease[1], Bhaskar Dasgupta[2], Richard Watts[3], Andrew Gough[4], John D Isaacs[5], Michael Green[6], Neil McHugh[7], Lesley Hordon[8], Sanjeet Kamath[9], Mohammed Nisar[10], Yusuf Patel[11], Chee-Seng Yee[12], Robert Stevens[12], Pradip Nandi[13], Anupama Nandagudi[14], Stephen Jarrett[15], Charles Li[16], Sarah Levy[17], Susan Mollan[18], Abdel Salih[19]

**Nursing and Study Team:** Oliver Wordsworth[1], Prisca Gondo[2], Jane Hollywood[2], Genessa Peters[3], Christine Routledge[5], Anne Gill[6], Lisa Carr[6], Rose Wood[7], Clare Williams[10], Mandy Oakley[10], Emma Sanders[11], Felicity Mackenzie[12], Rosanna Fong[12], Lynne James[13], Jenny Spimpolo[13], Andy Kempa[13], Karen Culfear[14], Asanka Nugaliyadde[14], Esme Roads[16], Bridie Rowbotham[18], Zahira Masqood[18]

[1]School of Medicine, University of Leeds and NIHR-Leeds Musculoskeletal Biomedical Research Unit, Leeds Teaching Hospitals NHS Trust, LS7 4SA, UK

[2]Southend Hospital, Prittlewell Chase, Westcliff-on-Sea, Essex, SS0 0RY, UK

[3]Ipswich Hospital, Heath Road, Ipswich, Suffolk, IP4 5PD, UK

[4]Harrogate District Hospital, Lancaster Park Rd, Harrogate, HG2 7SX, UK

[5]Freeman Hospital, Freeman Road, High Heaton, Newcastle upon Tyne, Tyne and Wear, NE7 7DN, UK

[6]York District Hospital, Wigginton Road, York, YO31 8HE, UK

[7]Royal National Hospital for Rheumatic Diseases, Upper Borough Walls, Bath, Avon, BA1 1RL, UK

[8]Dewsbury District and General Hospital, Halifax Road, Dewsbury, WF13 4HS, UK

[9]Haywood Hospital, High Lane, Burslem, Staffordshire, ST6 7AG, UK

[10]Queen’s Hospital, Belvedere Road, Burton-on-Trent, DE13 0RB, UK

[11]Hull Royal Infirmary, Anlaby Road, Hull, HU3 2JZ, UK

[12]Doncaster Royal Infirmary, Armthorpe Road, Doncaster, DN2 5LT, UK

[13]Northampton General Hospital, Cliftonville, Northampton, NN1 5BD, UK

[14]Basildon University Hospital, Nethermayne, Basildon, SS16 5NL, UK

[15]Pinderfields General Hospital, Aberford Road, Wakefield, WF1 4DG, UK

[16]Royal Surrey County Hospital, Egerton Road, Guildford, GU2 7XX, UK

[17]Croydon University Hospital, 530 London Road, Croydon, Surrey, CR7 7YE, UK

[18]Queen Elizabeth Hospital, Mindelsohn Way, Birmingham, B15 2TH, UK

[19]Warrington Hospital, Lovely Lane, Warrington, Cheshire, WA5 1QG, UK
